# EEG-Derived Proxies of Cortical Excitability in Epilepsy: Group Discrimination, Temporal Stability and Medication Sensitivity

**DOI:** 10.64898/2026.06.02.26354693

**Authors:** Gian Marco Duma, Natalia Valencia-Enciso, Javier Rasero, Paolo Bonanni, Giovanni Pellegrino

## Abstract

**Objective:** To identify EEG-derived markers of cortical excitability that best distinguish people with epilepsy from healthy controls while maintaining stability across repeated recordings, and to assess the influence of antiseizure medication (ASM) burden on these measures.

**Methods:** Scalp EEG recordings from the Temple University Hospital EEG Epilepsy Corpus were analyzed, including 1,404 recordings from 96 individuals with neurologist-confirmed epilepsy and 85 healthy controls across multiple sessions. Eight global EEG measures were evaluated: aperiodic exponent and offset, sample entropy, detrended fluctuation analysis exponent and derived index, spatial gamma-band phase consistency, and absolute and relative alpha power. Primary group inference used linear mixed-effects models to account for repeated recordings and sessions within participants. Secondary analyses examined group differences in measures temporal stability and cross-metric correlation. We examined only in epilepsy group associations between EEG-derived metrics and ASMs count, and their impact on cross-metric correlations.

**Results:** In the mixed-effects models, aperiodic offset and exponent were the only measures with FDR-robust group effects. Aperiodic offset showed higher consistency in epilepsy than in healthy controls (standardized epilepsy-minus-healthy beta = 0.52, 95% CI 0.25 to 0.78, FDR-adjusted p = 0.001). Across sessions, consistency was higher in epilepsy for aperiodic offset exponent (beta = 0.32, 95% CI 0.10 to 0.54, FDR-adjusted p = 0.018). Cross-metric correlation patterns were largely preserved between groups. Medication associations were examined separately within the epilepsy group. Higher ASM burden was associated with higher alpha relative power.

**Significance:** Among eight candidate EEG measures aperiodic offset and exponent differed between groups (epilepsy vs. healthy controls), and both were more reproducible across sessions in epilepsy. Relative alpha power tracked within-person antiseizure medication burden, suggesting that disease-related and treatment-related EEG markers may be complementary. These findings, obtained from routine clinical EEG in an open dataset, suggest aperiodic spectral parameters as candidates for prospective validation as biomarkers.

**Plain language Summary:** People with epilepsy show changes in brain activity that may reflect altered cortical excitability. We analyzed over 1,400 EEG recordings from people with epilepsy and healthy controls to compare several EEG-based excitability measures. Aperiodic EEG spectral measures were the most reliable markers of epilepsy and remained stable over time. Other measures were more sensitive to temporary brain states. Anti-seizure medications significantly influenced many EEG metrics and should be considered when interpreting EEG biomarkers.

**Key points:**

- Aperiodic offset and exponent were higher in epilepsy than in healthy controls.
- Aperiodic measures were more reproducible across sessions in epilepsy than controls.
- Within patients, more antiseizure medications were linked to higher relative alpha.
- The eight excitability proxies captured complementary, not interchangeable, dynamics.

## 1 Introduction

Epilepsy is a chronic disorder of brain excitability characterized by a persistent predisposition of neuronal networks to generate spontaneous, hypersynchronous discharges and seizures ^1^. Cortical excitability refers to the responsiveness of cortical neurons to incoming input, and is shaped by the dynamic balance between excitatory and inhibitory (E/I) neurotransmission, intrinsic membrane properties, and network-level interactions^2^. The central role of excitability in epilepsy is reflected in the mechanisms of antiseizure medications (ASMs). Although ASMs act through diverse molecular targets—including enhancement of GABAergic inhibition, attenuation of glutamatergic transmission, modulation of voltage-gated ion channels, and regulation of presynaptic neurotransmitter release—they converge on a common physiological effect: reducing pathological neuronal and network excitability, thereby limiting the emergence and propagation of epileptiform activity^3–6^.

Despite this well-established conceptual model, a reliable, practical, bedside measure of excitability has been lacking ^7^. Standard diagnostic tools, including electroencephalography (EEG) and neuroimaging, provide indirect markers of epileptogenicity but do not directly quantify the excitation–inhibition (E/I) balance in a clinically feasible and reproducible manner^3^. Perturbation approaches, including TMS-evoked responses in motor and nonmotor cortex, can probe aspects of cortical physiology in research settings ^2^. Although seizure induction is a recognized potential adverse event of TMS, its risk depends on stimulation parameters and individual clinical factors, and conventional protocols performed within established safety recommendations generally carry a low risk ^8,9^.

Recent works have proposed several complementary EEG-derived measures as candidate proxies of cortical excitability and related neural dynamics ^7,10–12^. These measures were selected because they capture distinct aspects of spontaneous neural activity that have been associated with excitation–inhibition balance, cortical physiology, or network organization ^13^. Moreover, given the routine use of EEG in the diagnosis and longitudinal monitoring of epilepsy, multiple measures hold potential for translation into potential clinical applications, enabling the assessment of excitability-related changes using a widely available, non-invasive recording modality. Candidate metrics include the aperiodic exponent and offset of the power spectral density, parameterized using FOOOF, which reflect the 1/f structure of neural activity and have been proposed as indirect markers of E/I balance ^14,15^. Additional measures include sample entropy, which quantifies signal complexity ^16,17^, detrended fluctuation analysis (DFA), which characterizes long-range temporal correlations^18^; spatial high-gamma phase consistency, reflecting large-scale network synchrony^19–21^, and alpha-band power, an established correlate of cortical inhibition ^22^. These measures should not be regarded as interchangeable or as direct measurements of cortical excitability or excitation–inhibition balance. In fact, a recent study from our group evidenced that the correlational structure of these metrics does not converge on a single dimension, emphasizing that they metrics capture multidimensional and non-overlapping properties of brain dynamics related to cortical excitability^13^. Alterations of these metrics in epilepsy were reported across multiple recording modalities, including scalp EEG ^3,20,23^, stereotactic EEG ^24^, and MEG ^22^. However, their relative performance, temporal stability, and robustness to clinical confounds have not been systematically evaluated within a common analytical framework ^25^.

A major unresolved question is whether these candidate resting state derived measures remain stable across repeated recordings within the same individual, a prerequisite for longitudinal monitoring ^26–29^. Routine clinical EEG is subject to substantial variability arising from artefacts, heterogeneous acquisition conditions, and changes in physiological state ^30–34^ ^35^. Furthermore, ASM treatment represents an important confound, as quantitative EEG measures can vary with medication load independently of underlying disease biology ^3,36–39^. Clinically useful metrics should be sensitivity to the clinical condition (i.e., epilepsy in this case) and should present reliability and stability across repeated recordings while remaining interpretable in the context of pharmacological treatment ^40^.

Here, we address these challenges using the open-access Temple University Hospital EEG Epilepsy Corpus (TUEP) dataset, which contains longitudinal scalp EEG recordings from individuals with epilepsy and controls, together with detailed clinical metadata, including ASM exposure at each session ^41,42^. The longitudinal structure of this dataset enables simultaneous assessment of inter-individual differences and intra-individual stability. Within epilepsy, the medication list allows an exploratory analysis using a coarse count of recognized ASM names, but metadata from the public repository do not quantify dose, duration, or treatment effect.

We compared complementary resting-state EEG features that have been proposed as candidate proxies of cortical physiology relevant to excitability. Specifically, we evaluate their group differences, temporal stability, and exploratory epilepsy-only associations with ASM count using a unified whole-brain analytical framework. Ultimately, this work compares candidate EEG-derived measures of cortical excitability, examining their potential relevance for characterizing epilepsy and monitoring changes across repeated recordings.

## 2 Materials and Methods

### 2.1 Data

Data were obtained from the TUEP dataset, an open-access repository of clinical scalp EEG recordings from individuals with epilepsy and from non-epilepsy controls^41,42^. Recordings were stored in European Data Format (EDF), and subjects could contribute multiple sessions acquired on different dates, enabling assessment of both between-subject differences and within-subject longitudinal stability.

Session-level metadata were extracted from the accompanying metadata file and linked to recordings using subject-session identifiers. The TUEP dataset does not provide structured patient-level diagnosis labels identifying epilepsy type and anatomical lateralization, preventing more detailed analysis on epilepsy type or spatial location of the effects. Medication names were parsed using a predefined dictionary of generic and brand antiseizure medications (ASMs), and the number of distinct ASMs prescribed at each recording was determined. Dose, daily schedule, treatment duration, adherence and serum concentration were not available, preventing more detailed analyses beyond those based on the number of ASMs.

### 2.2 EEG Preprocessing

All EEG recordings were processed in MATLAB using EEGLAB 2024.2.1^43^ and Brainstorm ^44^. Recordings were imported with pop_biosig, channel labels were standardized to the international 10–20 nomenclature, non-EEG channels and duplicate labels were removed, and electrode locations were assigned using the standard 10–05 template.

Segments corresponding to photic stimulation were automatically identified using stimulation channels and removed, including 10 s before the first pulse and 10 s after the last pulse. Recordings with fewer than 2 min of remaining data were excluded. Data sampled above 250 Hz were downsampled to 250 Hz, high-pass filtered at 1 Hz, and notch filtered between 59–61 Hz. Artifact removal was performed in two stages to preserve continuous EEG segments, as several measures are sensitive to signal discontinuities. An initial Artifact Subspace Reconstruction (ASR) pass was used only to generate a sample-level quality mask. The longest continuous clean segment was retained and classified as PASS_GOLD (≥5 min), PASS_MIN (2–5 min), or FAIL (<2 min); accepted segments were truncated to a maximum duration of 10 min. A second ASR pass was then applied to the selected segment using burst correction and Riemannian interpolation (ASR; clean_artifacts)^45^. Independent component analysis (ICA) was performed using extended Infomax. ICA weights were estimated on data downsampled to 100 Hz and transferred to the original sampling rate; principal component analysis was applied when necessary to compensate for rank deficiency. Components were classified using ICLabel^46^, and those representing eye, muscle, or cardiac artifacts were removed using probability thresholds (>0.80 artifact, <0.15 brain). Channels rejected during preprocessing were reconstructed using spherical spline interpolation. All recordings underwent visual quality control based on their power spectra before being retained for analysis.

### 2.3 Measures

For each cleaned recording, we computed a set of complementary EEG-derived proxies of cortical excitability using the implementation described in Pellegrino et al.^7^. All measures were first computed at the channel level and then averaged across channels to obtain a single global value per recording. The measures were selected because prior work has related them to neural activity and excitability-related physiology; in the present passive resting-state data, they are treated as candidate proxies rather than direct physiological measurements ^13^

The analyzed measures were:

i. the aperiodic exponent (Exponent in figures) and aperiodic offset (Offset in figures) of the power spectral density ^15^;
ii. sample entropy ^16,17,47^ (Entropy in figures);
iii. the detrended fluctuation analysis (DFA exponent in figures) scaling exponent of the alpha-band amplitude envelope, together with the derived fE/I index (fE/I in figures) ^18^;
iv. spatial phase consistency in the high-gamma band (30–59 Hz, Gamma Synchrony index in figures), calculated as the phase-locking value averaged across all channel pairs ^21,31,48,49^_;_
v. absolute and relative alpha power in the 8–13 Hz band (AbsoluteAlpha and AlphaRelative in figures, respectively) ^22,50^.

Based on prior work, lower aperiodic exponent and lower alpha power were interpreted as reflecting relatively greater excitation, whereas higher entropy, higher DFA-related measures, and higher gamma-band spatial consistency were interpreted as reflecting greater cortical or network excitability. See Supplementary Materials for the computational details. Descriptive statistics of the sample and the measure are reported in the Supplementary Material (Table S1, S2)

### 2.4 Statistical analysis

All analyses were performed in MATLAB.

#### Group differences

For each measure, we fitted a linear mixed-effects model with group as a fixed effect and random intercepts for participant and for session nested within participant: measure ∼ group + (1 | participant) + (1 | participant-session). This accounts for repeated recordings per participant and for correlation among recordings from the same session. Healthy participants were the reference group. Models including age, and age and sex, were run as sensitivity analyses.

#### Temporal stability

Within-participant consistency was estimated separately in each group with nested variance-components models (measure ∼ 1 + (1 | participant) + (1 | participant-session)). From these we derived two intraclass correlations (ICC): same-session consistency (recordings from the same participant and session) and across-session consistency (recordings from the same participant in different sessions). Group differences in ICC were tested with participant-level permutation, and confidence intervals were obtained by participant-level bootstrap.

#### Cross-measure structure

We compared the Spearman correlation matrices of the eight measures between healthy and epilepsy participants using a global permutation test on the difference between matrices, with individual pairs tested descriptively. This was repeated at recording, session and participant level to check robustness to aggregation.

#### Medication

For each participant-session, ASM exposure was the number of distinct ASMs. We split it into a within-person component (session count minus the participant’s mean count) and a between-person component (participant mean count) and fitted, for each measure, session-mean measure ∼ within-person ASM count + participant mean ASM count + (1 | participant). The within-person coefficient estimates the change in a measure associated with one additional ASM relative to the participant’s usual regimen. The between-person coefficient is cross-sectional and may be confounded by indication, severity or clinical history. For the analysis details see Supplementary Materials (S2 section).

## 3 Results

### 3.1 Dataset overview

After preprocessing and aggregation, the analyses were performed at three levels: 1,404 recordings (208 healthy and 1,196 epilepsy), 575 sessions (133 healthy and 442 epilepsy), and 181 subject-level averages (85 healthy and 96 epilepsy). Medication analyses were based on the 442 epilepsy sessions with available ASM information. For the cross-correlation analysis stratified by medication burden, 251 epilepsy sessions fell in the lower-burden group (0–1 ASM) and 191 in the higher-burden group (≥2 ASMs).

### 3.2 Group differences in EEG-derived excitability measures

After false-discovery-rate correction, aperiodic offset was higher in epilepsy than in healthy controls (epilepsy-minus-healthy difference, .263 native units; 95% CI: [.099 - .427]; *p*_adjusted_ = .014). Aperiodic exponent was also higher in epilepsy (difference, .184; 95% CI: [.046 - .323]; *p*_adjusted_ = .037). Absolute alpha power showed a nominal group difference (*p* = .021) that did not survive FDR correction (*p*_adjusted_ = .055). No other measure showed a group difference (all *p*_adjusted_ > .05) (see Fig.1; Table S3). When investigating the more complex model with Age and Sex as covariates, we observed that only aperiodic offset met the FDR threshold in all three specifications. Aperiodic exponent met the FDR threshold in the group-only model, but not after adding age, with or without sex. The direction and magnitude of the exponent coefficient remained similar across models (Figure S1).

**Figure 1.**
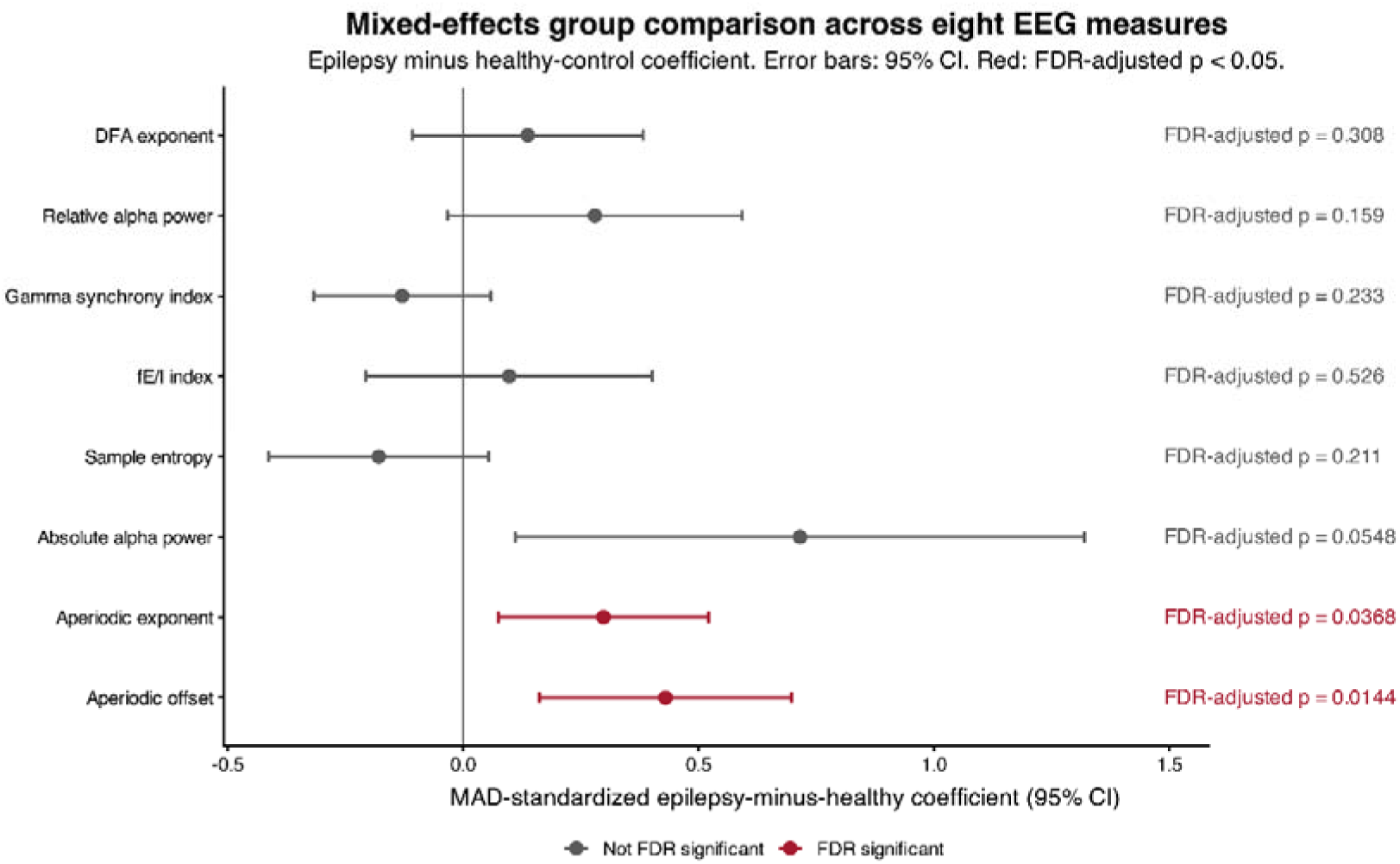
Mixed-effects group comparison across eight candidate resting-state EEG measures. Points show the epilepsy-minus-healthy group coefficient and horizontal bars show 95% confidence intervals. Each measure was analyzed using a linear mixed-effects model with a fixed group effect and random intercepts for participant and participant-session nested within participant. Healthy participants were the reference group. Red indicates *p*_adjusted_ < .05 across eight measures. Coefficients are median absolute deviation (MAD)-standardized only for cross-measure visualization.

### 3.3 Consistency across repeated recordings and sessions

Within-session consistency differed between groups for two measures after FDR correction. Aperiodic offset showed higher consistency in epilepsy than in healthy controls (intraclass correlation [ICC], 0.80 vs 0.21; difference, 0.59; 95% bootstrap CI: [−.004 - 0.80]; *p*_adjusted_ = .008), as did absolute alpha power (ICC, 0.87 vs 0.32; difference, 0.55; 95% bootstrap CI, [−.09 - 0.70]; *p*_adjusted_ = .008). Across sessions, consistency was higher in epilepsy for aperiodic offset (ICC, 0.47 vs 0.001; difference, 0.47; 95% bootstrap CI, 0.08 to 0.55; *p*_adjusted_ = .008) and aperiodic exponent (ICC, 0.29 vs 0.02; difference, 0.28; 95% bootstrap CI, −0.03 to 0.35; *p*_adjusted_ = .008). No other healthy-versus-epilepsy consistency difference survived FDR correction (see Fig.2A-B).

**Figure 2.**
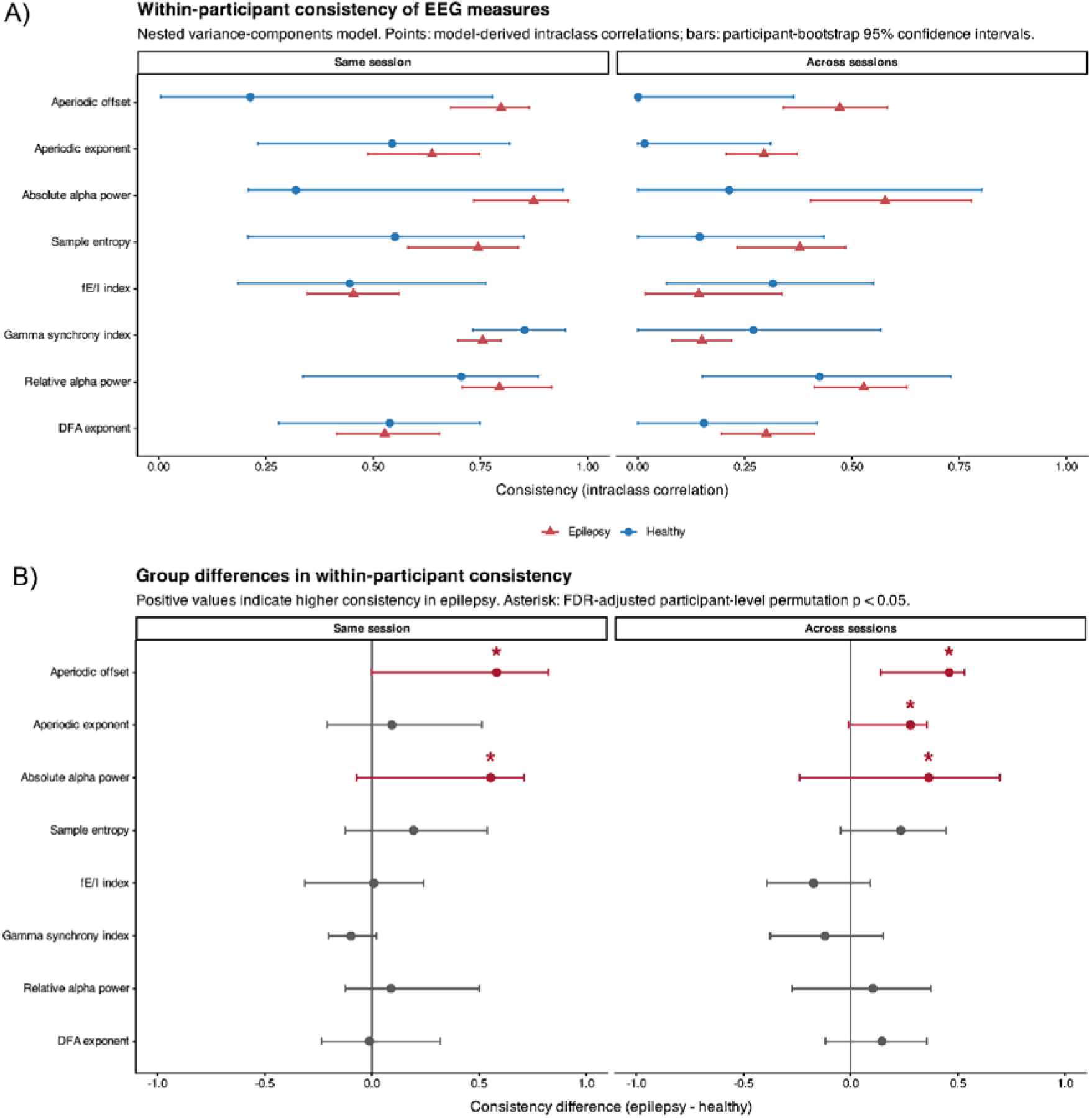
Within-participant consistency of eight candidate EEG measures in healthy participants and participants with epilepsy. In panel A, points show model-derived intraclass correlations from nested variance-components models, and bars show participant-bootstrap 95% confidence intervals. Same-session consistency is the expected correlation between two recordings from the same participant and session. Across-session consistency is the expected correlation between recordings from the same participant obtained in different sessions. Panel B shows healthy-versus-epilepsy differences in within-participant consistency. Points show epilepsy-minus-healthy differences in intraclass correlation and bars show participant-bootstrap 95% confidence intervals. Positive values indicate higher consistency in epilepsy. Asterisks indicate < .05 from participant-level group-label permutation tests, corrected across eight measures separately for same-session and across-session endpoints.

### 3.4 Cross-correlation across measures

Cross-correlation analysis was used to assess whether the candidate excitability markers captured a shared or reorganized physiological structure across groups.

Overall, the inter-measure correlation pattern was broadly similar in healthy controls and (Fig.3). The primary participant-level correlation matrices did not differ significantly between groups (global Fisher-z squared distance = .844; *p*_adjusted_ = .552 across the three aggregation levels). No individual cross-measure correlation difference survived FDR correction across the 28 pairwise tests. Thus, this analysis found no evidence for a broad group difference in the covariance pattern among the eight candidate EEG measures. The conclusion was consistent across all aggregation levels: recording-level *p*_adjusted_ = .552, session-level *p*_adjusted_ = .628, and participant-level *p*_adjusted_ = .552. The visual patterns of positive and negative associations were broadly similar in the two groups and were not materially changed by whether repeated recordings were retained, averaged within sessions, or averaged within participants (Figure S2).

**Figure 3.**
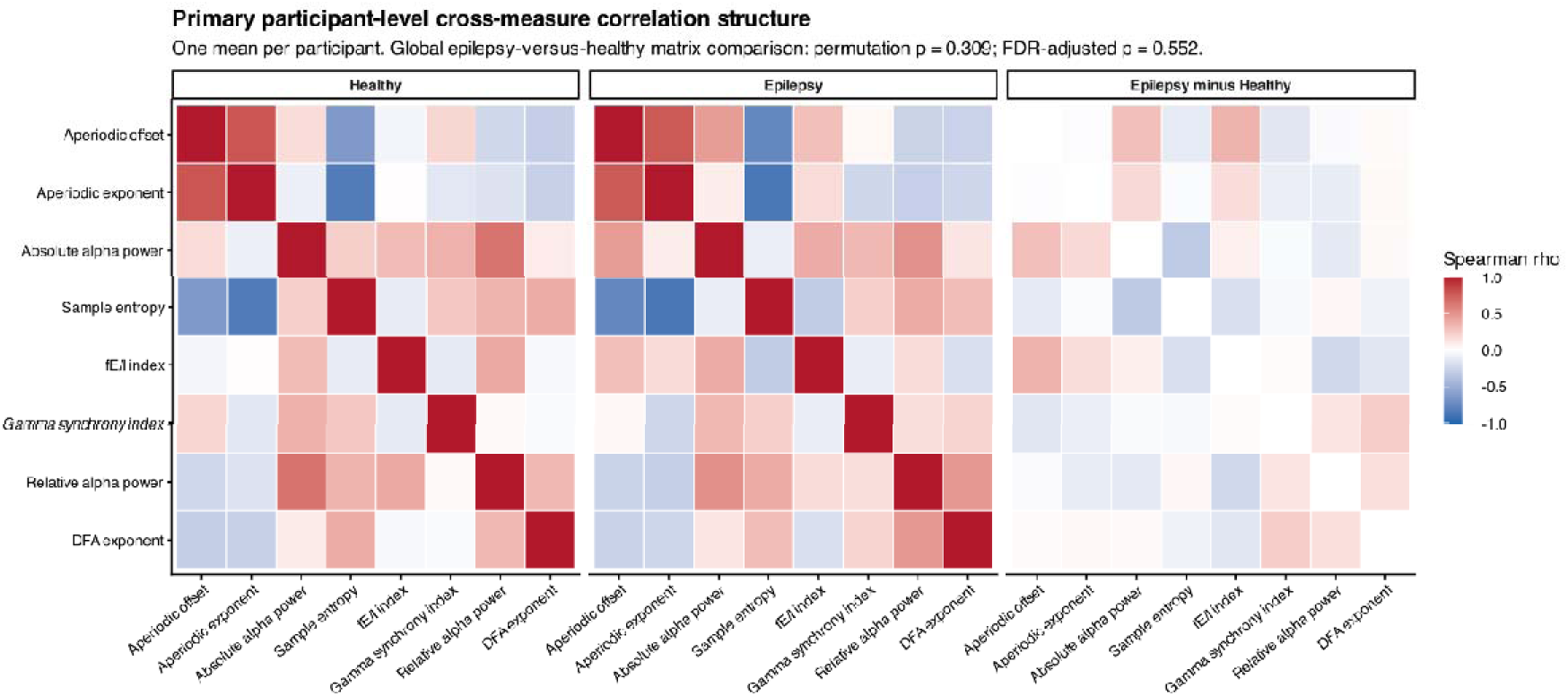
Primary participant-level cross-measure correlation structure. Each participant contributes one mean value per measure. Heatmaps show Spearman correlations in healthy participants, epilepsy participants, and their difference (epilepsy minus healthy). The global difference between matrices was not significant (permutation p = 0.309; FDR-adjusted p = 0.552).

### 3.5 Medication effects

After FDR correction, relative alpha power was the only measure associated with within-person ASM-count change. Relative alpha power was higher by .0082 per additional ASM relative to the participant’s own average (95% CI: [.0040 - .0124]; *p_adjusted_* = .001). Offset, entropy, and DFA had nominal within-person associations (raw p < 0.05) that did not survive FDR correction. No between-person mean ASM-count association survived FDR correction (Fig.4; TableS4). No within-person ASM-count association with within-session variability or across-session deviation survived FDR correction across the eight measures. Thus, these exploratory analyses did not provide evidence that session-to-session changes in ASM count were associated with a systematic change in the short-term or across-session variability of these measures (Figure S3). The descriptive medication-category correlation matrices showed broadly similar patterns of co-variation across ASM-count categories. No formal inferential conclusion about a medication effect on correlation structure is made because categories overlap within participants and there are no exposure-randomized comparisons (Figure S4).

**Figure 4.**
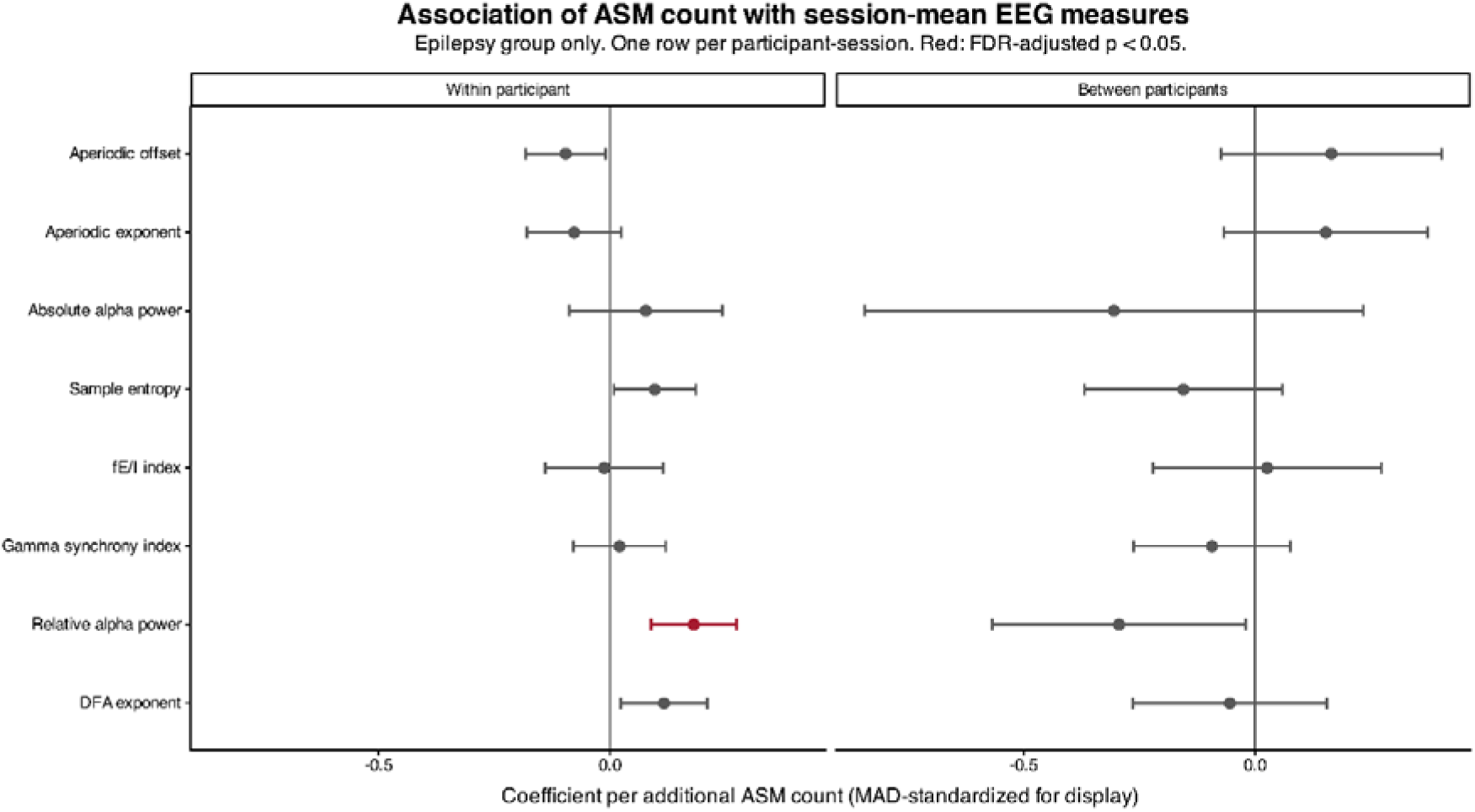
Association of ASM count with session-mean EEG measures. The within-person panel is the primary medication analysis; it compares each participant’s sessions with their own average ASM count. The between-person panel is cross-sectional and is more susceptible to confounding by indication. Red indicates FDR-adjusted p < 0.05. Effect sizes are MAD-standardized only for common visual scaling; all inferential estimates in Tables 1 and 2 are in native units.

## 4 Discussion

In this study, we performed a systematic, large-scale evaluation of EEG-derived proxies of cortical excitability in epilepsy, leveraging the longitudinal structure of the Epilepsy Corpus EEG open database. By comparing multiple candidate metrics within a unified analytical framework, we sought to determine their: i) performance in terms of group (epilepsy vs healthy) differences; ii) temporal stability; iii) cross-metric consistency and iv) examine the influence of medication within the epilepsy group. Several key findings emerged from this analysis.

### 4.1 Group differences

Aperiodic spectral parameters (offset and exponent) were the only EEG-derived metric showing group differences after correction for multiple comparisons in the mixed-effect models. Specifically, patients displayed a larger aperiodic offset and exponent values as compared to controls. The higher aperiodic exponent and offset in epilepsy agree with previous scalp and intracranial work, including our own, in focal epilepsy ^3,23,52^. Within the E/I framework, a steeper spectral slope is usually read as relatively stronger inhibition ^14,15^. However, this interpretation is consistent with previous work from our group and others showing increased inhibitory tone during the interictal and peri-ictal periods ^3,23,24,53^. In this context, the findings align with the interictal suppression hypothesis, according to which epileptic networks recruit inhibitory processes during the interictal state to constrain pathological synchronization and reduce seizure likelihood. Under this view, the elevated aperiodic exponent may reflect a relatively stable background state of compensatory regulation that coexists with an underlying susceptibility to seizure generation. Other studies on invasive recordings do not confirm the findings above, but still advocate for aperiodic properties to be considered excitation / inhibition markers, and to fluctuate overtime ^52^. While findings in focal epilepsy (including TLE) consistently show higher exponent suggesting inhibitory compensation, studies of generalized epilepsy have found the opposite, lower exponent consistent with hyperexcitability ^54^. The composition of the current cohort may account for this directionality. Additionally, the persistence of these effects after averaging across subjects suggests that aperiodic parameters reflect a relatively trait-like property of epileptic networks rather than transient state fluctuations ^26,35^. Because the database has no structured syndrome or lateralization labels, we could not test whether the direction of the effect depends on epilepsy type. A heterogeneous, probably focal-dominated cohort may therefore explain the direction we observed. Two caveats limit how far these findings can be interpreted. First, the exponent effect was less robust than the offset effect. It met the FDR threshold in the group-only model but not after adjusting for age, with or without sex, although its direction and magnitude were similar. The offset effect held in all three specifications. We therefore consider the offset the more reliable group marker here and the exponent a consistent but more tentative one. Second, the aperiodic exponent and offset are correlated descriptors of the same spectral fit. Broadband offset is sensitive to overall signal amplitude, which can be influenced by non-neural factors such as skull and scalp properties. The remaining six measures did not show a group difference after correction. Their sensitivity to epilepsy in this dataset is therefore not supported, and we cannot rule out smaller effects that this global, heterogeneous routine-EEG approach was not able to detect.

### 4.2 Temporal stability

In epilepsy, aperiodic offset and exponent were reproducible across sessions acquired on different days, whereas in healthy controls they were essentially not reproducible (ICC ≈ 0). We interpret this as a difference in between-subject variance. In healthy adults the aperiodic spectrum varies little between people compared with its fluctuations with vigilance, arousal and recording conditions, so the ICC is low. In epilepsy, patients differ markedly in etiology, network involvement and treatment, and these differences appear to be expressed in a lasting individual spectral profile. This fits the view that elevated aperiodic offset and exponent reflect a chronic, trait-like property of epileptic networks (for example a persistent compensatory inhibitory tone) rather than a transient interictal state, and it links the two main findings of the study. Alpha power followed a different pattern. Absolute alpha was highly consistent within a session in epilepsy (ICC 0.87) but not across sessions. Alpha is strongly modulated by vigilance, drowsiness, eye state and attention, which are largely constant within a recording but differ between visits, as this pattern would predict. Together, these results suggest that different measures may serve different purposes. Aperiodic parameters could provide a stable individual baseline against which changes, for example after a treatment change or before a seizure, are more easily detected, whereas alpha-based measures may better capture the current functional state, in line with their sensitivity to medication (see 4.4).

### 4.3 Cross-metric consistency

The overall correlation structure among measures was largely preserved between epilepsy and controls. This indicates that these candidate proxies, while capturing distinct aspects of neural dynamics, maintain a broadly similar pattern of interrelationships across groups ^13^. In other words, epilepsy does not fundamentally rewire the way these measures co-vary; rather, it appears to displace the network along a shared axis of excitability without generating a qualitatively different pattern of interdependencies. All the measures tested here were selected as they are believed to reflect the excitation-inhibition balance, and as such one would expect them to co-vary, or at least vary in the same direction. Yet, they often diverged. No single metric fully captures cortical excitability. Rather, different measures appear to reflect complementary dimensions of neural dynamics, suggesting to not treat any one of them as a stand-alone index of E/I balance.

### 4.4 Medication effects

The most consistent medication finding was that, within the same person, sessions with one additional ASM showed higher relative alpha power. We interpret this as a signature of the shift toward inhibition that ASMs are designed to produce. Most ASMs, through enhanced GABAergic transmission, reduced glutamatergic transmission or modulation of voltage-gated channels, reduce network excitability ^6^, and alpha-band activity is considered a correlate of cortical inhibition and of the thalamocortical gating of cortical activity^55^ . A higher relative alpha with more drugs is therefore in the direction expected if medication increases inhibitory tone and stabilizes cortical dynamics. The fact that the effect was seen in relative rather than absolute alpha suggests a redistribution of spectral power toward the alpha band, and not simply a global change in amplitude. The within-person design strengthens this interpretation, since each patient serves as their own control and stable differences in epilepsy type, etiology and severity are removed. The between-person comparison did not show this association, and this is also informative. Patients receiving more drugs generally have more severe or refractory epilepsy, whose EEG may differ from that of milder cases in the opposite direction. The disease-related and drug-related contributions to alpha power may therefore partly offset each other when patients are compared with one another, but become visible when each patient is compared with themselves. Finally, changes in ASM count were not associated with changes in within-session or across-session variability. A reasonable interpretation is that medication shifts the level of these measures without altering how reproducible they are, so that each patient retains a characteristic profile under treatment. This is consistent with the idea that the stable aperiodic signature persists across treatment changes. Because exposure was a coarse count that pooled drugs with different mechanisms, and because changes in treatment follow clinical course, these associations should be seen as pointing to a general shift toward inhibition and not as drug-specific effects. More granular data (e.g., daily dosage) would allow a mechanistic test of this account.

### 4.4 Translational value and possible applications

These results do not support using any of the evaluated measures as a stand-alone diagnostic biomarker. The aperiodic parameters were the most promising candidates, and their detectability in routine, heterogeneous clinical EEG is encouraging. The offset and exponent are better viewed as candidates for further validation, for example in prospective cohorts with defined epilepsy types, drug-naïve patients and standardized acquisition. Our findings on relative alpha power suggest that medication status should be recorded and modelled whenever such measures are considered for treatment monitoring.

### 4.5 Limitations and future directions

Several limitations should be acknowledged. First, the use of routine clinical EEG introduces variability in recording conditions, electrode configurations, and patient state, all of which may influence the measured signals. Although this heterogeneity enhances generalizability, it may also reduce sensitivity to subtle effects. Second, our analysis adopted a global, whole-brain approach and therefore does not account for spatial heterogeneity or focal epileptogenic regions. This limitation was intrinsically related to the available data. In fact, TUEP dataset does not contain ready to use information regarding the epilepsy type (e.g., focal vs generalized), nor location distinction in case of focal epilepsy. This prevents inference on the significance of potential electrode based analysis. Third, ASM burden was quantified as the number of medications rather than dosage or pharmacological class, limiting the precision of medication-related analyses. Finally, the observational nature of the dataset precludes causal inference regarding the relationships among excitability measures, epilepsy, and treatment.

Future work should aim to integrate spatially resolved measures, incorporate more detailed pharmacological information, and validate these findings in prospective, standardized datasets. Combining EEG-derived metrics with other modalities, such as structural or functional neuroimaging, may further improve characterization of epileptic networks. In addition, the development of composite indices of excitability, potentially informed by machine learning approaches, represents a promising avenue for translating these findings into clinically actionable tools.

## 5 Conclusion

In summary, this study provides a comprehensive evaluation of EEG-derived proxies of cortical excitability in epilepsy. Our findings highlight the robustness of aperiodic spectral parameters, and the influence of ASM treatment on electrophysiological signatures. These findings support aperiodic parameters as candidates for further investigation, and they show that multiple, complementary measures and careful accounting for medication are needed in studies of EEG-based excitability markers. Our results also confirm that quantitative analysis of excitability markers extracted from routing recordings carry relevant information on epilepsy.

## Data availability statement

The data used for this study are already accessible in a open data repository: https://isip.piconepress.com/projects/nedc/html/tuh_eeg/index.shtml

## Patient consent

The study uses open available data from which the consent was already been acquired.

