## Supplementary Material for "EEG-Derived Proxies of Cortical Excitability in Epilepsy: Group Discrimination, Temporal Stability and Medication Sensitivity"

Supplementary Materials

**Additional Methods:**

**Segments data specification**

Each accepted recording contributed one contiguous clean segment. See Supplementary material for the details about segments duration across groups The preprocessing pipeline targeted 5 min of clean data, required at least 2 min, and truncated accepted segments at 10 min. We applied offline average re-referencing after artifact cleaning and channel interpolation. Retained segment duration was calculated as the number of samples in F divided by the sampling rate (250 Hz). Across 181 participants (96 epilepsy, 85 healthy), the median duration was 311.6 s (IQR 195.8-589.6 s; range 120.1-600.0 s). More in detail, the median was 320.1 s (IQR 202.1-590.9 s) for 1,196 epilepsy recordings and 284.6 s (IQR 167.5-567.7 s) for 208 healthy recordings. At the participant level, the median number of retained segments was 3.5 (range 1-263) in epilepsy and 1.0 (range 1-18) in healthy participants; the corresponding median total retained durations were 19.7 min (range 2.0-1,948.5 min) and 9.0 min (range 2.0-109.3 min), respectively.

**S2 Statistical analyses**

Mixed Model: For each EEG measure, we fitted the linear mixed-effects model used for the primary analysis in the manuscript, adding Gender and Sex as covariates with random intercepts for participant and session within participant. We obtained tree models where were later compared:

model1: measure ~ group + (1 | participant) + (1 | participant-session)

model2: measure ~ group + Age + (1 | participant) + (1 | participant-session)

model3: measure ~ group + Age + Sex + (1 | participant) + (1 | participant-session)

Age was centered at the sample mean (58.5 years), and female was the reference category for sex. Group effects were evaluated across the eight measures using FDR correction within each model specification. Healthy participants were older on average than participants with epilepsy, whereas the sex distribution was similar between groups (Table S1). Aperiodic offset met the FDR threshold in all three specifications. Aperiodic exponent met the FDR threshold in the group-only model, but not after adding age, with or without sex. The direction and magnitude of the exponent coefficient remained similar across models. None of the other six measures met the FDR threshold in any specification. Adding sex produced little additional change beyond the age-adjusted model (Figure S1).

Consistency analysis. The variance-components models partitioned variability into participant, session-within-participant and recording-level residual components. Same-session consistency is the expected correlation between two recordings from the same participant and session, and across-session consistency is the expected correlation between recordings from the same participant in different sessions. Confidence intervals were obtained by resampling participants with all of their sessions and recordings retained (1000 bootstrap samples per group). Healthy-versus-epilepsy differences were tested by permuting group labels at the participant level (1000 permutations), preserving the complete nested structure of each participant. P-values were FDR-corrected across the eight measures, separately for same-session and across-session consistency.

Cross-measure correlation analysis. Participant-level analysis: all recordings were averaged within participants to give one value per measure per participant. Spearman correlation matrices were computed separately for healthy participants and participants with epilepsy. The primary global comparison summarized the difference between the two 8 × 8 matrices as the sum of squared Fisher-z-transformed differences across the 28 unique measure pairs. Group labels were permuted at the participant level (1000 permutations) to obtain a global p-value. As a secondary descriptive analysis, each of the 28 pairwise differences was tested with participant-level permutations and FDR-corrected across the 28 tests. Aggregation robustness: the same group-matrix comparison was repeated at the recording level (each accepted recording contributed), the session level (recordings first averaged within participant-session) and the participant level (recordings averaged within participant). Recording- and session-level correlations were participant-balanced so that participants contributing more observations did not dominate the estimates. The global test used participant-label permutation, preserving each participant’s repeated-measure profile, and the three global p-values were FDR-corrected across aggregation levels.

Medication analysis. All recordings were averaged within each participant-session so that the EEG outcome and medication exposure were aligned at the same level. ASM count was decomposed into a within-person component (session ASM count minus the participant’s mean ASM count) and a between-person component (participant’s mean ASM count across sessions). P-values were FDR-corrected across the eight measures, separately for the within- and between-person terms.

**Excitability Indices computational details:**

**The analyzed measures were:**

(i) the aperiodic exponent (exponent in figures) and aperiodic offset (Offset in figures) of the power spectral density, estimated with Brainstorm's built-in MATLAB FOOOF process using its default configuration: 1-40 Hz fitting range; fixed aperiodic mode; Gaussian peak model; peak-width limits of 0.5-12 Hz; maximum three peaks; 3 dB minimum peak height; proximity threshold of 2 standard deviations; no ignored line frequencies; and no guess-weight penalty;

(ii) sample entropy, as a measure of signal irregularity. Entropy measures have been linked to E/I. Lower entropy is linked to higher E/I, whereas higher entropy is linked to lower E/I (Entropy in figures). We computed Sample Entropy by including the code of Cannard [60] in the Brainstorm toolbox.

(iii) the detrended fluctuation analysis (DFA exponent in Figures) scaling exponent and the derived fE/I index (fE/I in figures). Within each selected frequency band, signals were filtered in consecutive 1-Hz bins and transformed to Hilbert amplitude envelopes. DFA was the slope of log10 mean detrended fluctuation versus log10 window size after integrating the mean-centered envelope; 50% overlapping windows were used. fE/I used 5-s windows with 80% overlap and was calculated as 1 minus the Pearson correlation between window-level normalized detrended fluctuations and window-level mean amplitude. fE/I values were set to missing when the corresponding DFA exponent was <=0.6 before output handling; adapting code from here https://github.com/annevannifterick/fEI_in_AD;

(iv) Global phase synchrony was quantified using the Meisel global order parameter, calculated at each time point as:

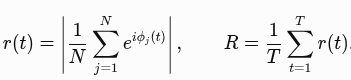

where (N) is the number of included channels and (*ϕ_j_(t)*) is the instantaneous phase of channel (*j*). The resulting time series was then averaged over time to obtain the global synchrony index (R).

Originally proposed and validated in epilepsy populations, this index quantifies the mean spatial phase synchronization across channels, particularly in the high-gamma frequency range (23,24). Previous studies have demonstrated a strong association between this measure and cortical excitability, as assessed using perturbational methods and its modulation by antiseizure medications (ASMs) in patients with epilepsy (16,23,24). Conceptually, this measure is related to the widely used phase-locking value (PLV), but differs in that it quantifies phase coherence across spatially distributed channels at each time point, rather than estimating phase locking between pairs of signals over time. Following previous recommendations, we focused on the high-gamma band (30–59 Hz). Importantly, this index is interpreted as a global, excitability-related measure of spatial phase synchrony rather than a measure of pairwise functional connectivity. It may capture zero-lag synchronization arising from shared signals or volume conduction/field spread and therefore does not, on its own, establish physiological connectivity between cortical regions.

(v) absolute and relative alpha power in the 8–13 Hz band (AbsoluteAlpha and AlphaRelative in figures, respectively): We performed Welch decomposition to compute the power spectrum density (PSD). We successively applied the spectrum renormalization based on relative power. Alpha power was extracted both for absolute and relative power spectrum as the sum of the power between 8-13Hz.

**Additional Results**

This supplement provides complete descriptive and inferential results. Raw-scale summaries are descriptive; primary case-control inference is based on the recording-level mixed-effects models reported in Supplementary Table S3. All statistical results are reproduced from saved analysis outputs.

### Table S1. Dataset composition and participant contribution

| **Group** | **Participants** | **Sessions** | **Recordings** | **Recordings/participant median [Q1, Q3]** | **Range** | **Sessions/participant median [Q1, Q3]** | **Range** |
| --- | --- | --- | --- | --- | --- | --- | --- |
| Healthy | 85 | 133 | 208 | 1.0 [1.0, 2.0] | 1.0 to 18.0 | 1.0 [1.0, 2.0] | 1.0 to 9.0 |
| Epilepsy | 96 | 442 | 1196 | 3.5 [1.0, 12.2] | 1.0 to 263.0 | 3.0 [1.0, 6.0] | 1.0 to 29.0 |

### Table S2. Participant-level raw-scale descriptive statistics by group

| **Measure** | **Group** | **Unit/scale** | **n** | **Median** | **Q1** | **Q3** | **Mean** | **SD** | **Min** | **Max** |
| --- | --- | --- | --- | --- | --- | --- | --- | --- | --- | --- |
| AbsoluteAlpha | Healthy | Native metric units* | 85 | 2.497e-12 | 1.258e-12 | 4.455e-12 | 4.004e-12 | 4.300e-12 | 1.186e-13 | 1.909e-11 |
| AbsoluteAlpha | Epilepsy | Native metric units* | 96 | 3.695e-12 | 1.800e-12 | 6.161e-12 | 5.442e-12 | 6.228e-12 | 2.732e-13 | 4.258e-11 |
| AlphaRelative | Healthy | Native metric units* | 85 | 0.0809 | 0.0412 | 0.1170 | 0.0875 | 0.0593 | 0.0056 | 0.3044 |
| AlphaRelative | Epilepsy | Native metric units* | 96 | 0.0764 | 0.0503 | 0.1246 | 0.0902 | 0.0504 | 0.0155 | 0.2305 |
| fE/I | Healthy | Native metric units* | 85 | 0.8047 | 0.7710 | 0.8618 | 0.8140 | 0.0653 | 0.6900 | 1.0235 |
| fE/I | Epilepsy | Native metric units* | 96 | 0.8055 | 0.7744 | 0.8492 | 0.8135 | 0.0510 | 0.7016 | 0.9791 |
| DFA exponent | Healthy | Native metric units* | 85 | 0.9380 | 0.8471 | 1.0063 | 0.9293 | 0.1533 | 0.5805 | 1.4942 |
| DFA exponent | Epilepsy | Native metric units* | 96 | 0.9481 | 0.8726 | 1.0081 | 0.9354 | 0.1317 | 0.3970 | 1.2136 |
| Entropy | Healthy | Native metric units* | 85 | 1.6644 | 1.3750 | 1.8851 | 1.6264 | 0.3608 | 0.7226 | 2.2563 |
| Entropy | Epilepsy | Native metric units* | 96 | 1.5399 | 1.3658 | 1.7686 | 1.5370 | 0.3307 | 0.5424 | 2.2250 |
| Gamma Syncrhony Index | Healthy | Native metric units* | 85 | 0.6101 | 0.4356 | 0.7507 | 0.5945 | 0.1711 | 0.3016 | 0.9044 |
| Gamma Syncrhony Index | Epilepsy | Native metric units* | 96 | 0.5754 | 0.4776 | 0.6583 | 0.5848 | 0.1288 | 0.3413 | 0.9242 |
| exponent | Healthy | Native metric units* | 85 | 1.2733 | 1.0039 | 1.5718 | 1.3125 | 0.4676 | 0.2713 | 2.6053 |
| exponent | Epilepsy | Native metric units* | 96 | 1.4769 | 1.2315 | 1.7445 | 1.5041 | 0.4013 | 0.6622 | 2.4530 |
| Offset | Healthy | Native metric units* | 85 | -10.8607 | -11.1515 | -10.4921 | -10.8400 | 0.5243 | -12.1586 | -9.8476 |
| Offset | Epilepsy | Native metric units* | 96 | -10.6094 | -10.9285 | -10.2490 | -10.5645 | 0.5425 | -11.7608 | -9.0300 |

*Note. One row per participant after averaging available recordings and sessions. These raw-scale summaries are descriptive and are not independent-observation tests. *Native metric units; unit not recoverable from retained materials. This applies to all metrics, including absolute alpha power.*

**Table S3. Primary mixed-effects group models**

| Measure | Epilepsy minus healthy | 95% CI | P | FDR-adjusted P | Conclusion |
| --- | --- | --- | --- | --- | --- |
| Aperiodic offset | 0.263 | 0.099 to 0.427 | 0.0018 | 0.014 | FDR significant |
| Aperiodic exponent | 0.184 | 0.046 to 0.323 | 0.0092 | 0.037 | FDR significant |
| Absolute alpha power | 1.90 x 10^-12 | 2.96 x 10^-13 to 3.50 x 10^-12 | 0.0205 | 0.055 | Nominal only |
| Sample entropy | -0.081 | -0.186 to 0.025 | 0.132 | 0.211 | Not significant |
| fE/I index | 0.013 | -0.028 to 0.054 | 0.526 | 0.526 | Not significant |
| Gamma synchrony index | -0.028 | -0.068 to 0.012 | 0.175 | 0.233 | Not significant |
| Relative alpha power | 0.014 | -0.002 to 0.029 | 0.079 | 0.159 | Not significant |
| DFA exponent | 0.010 | -0.007 to 0.027 | 0.269 | 0.308 | Not significant |

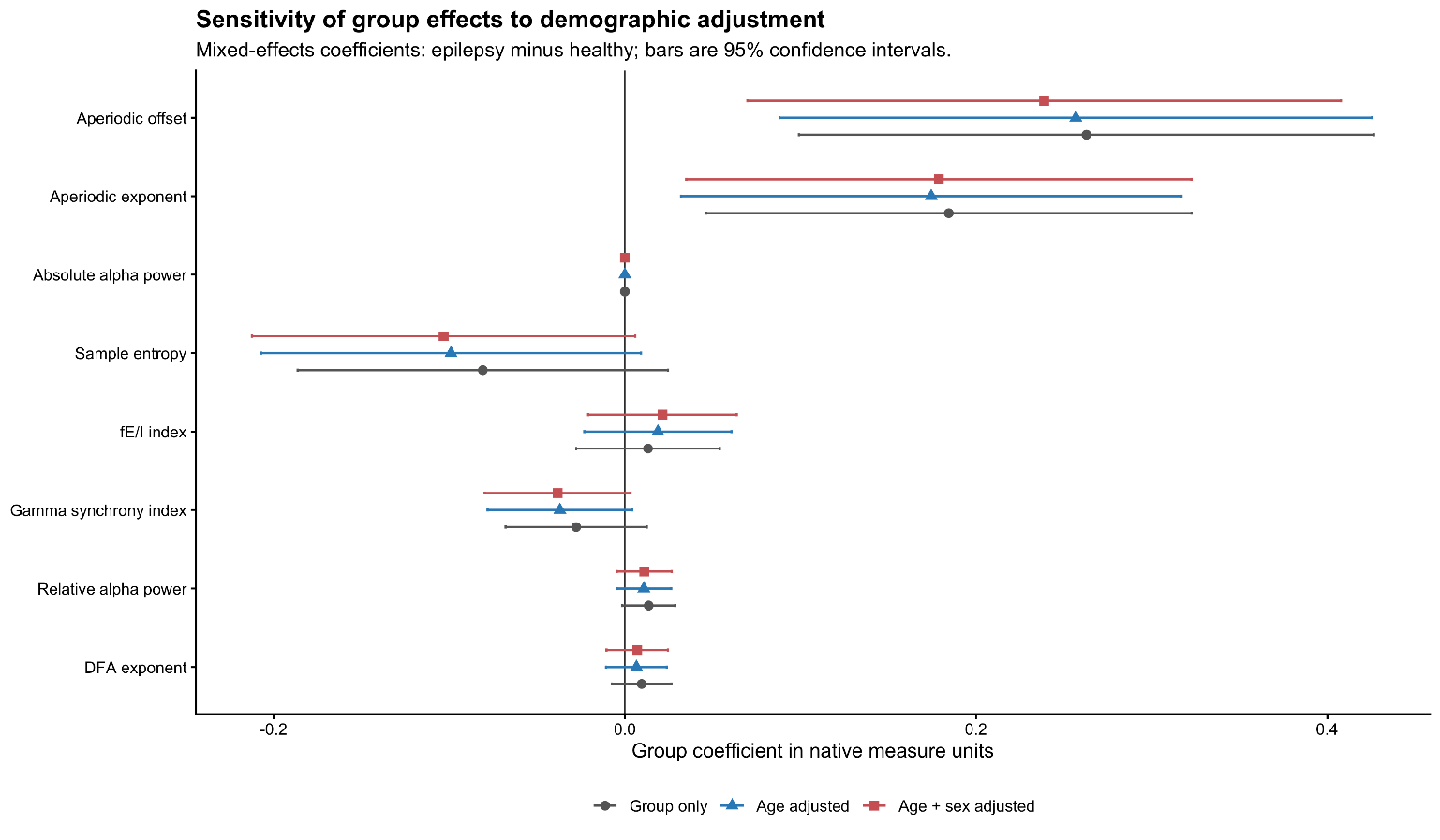

**Figure S1. Mixed-effects models comparison.** Points show the epilepsy-minus-healthy group coefficient and horizontal bars show 95% confidence intervals. Each measure was analyzed using a linear mixed-effects model with a fixed group effect and random intercepts for participant and participant-session nested within participant. The figure represents the comparison of the group effects for each EEG-derived metric measures across the three models. Red indicates FDR-adjusted P<0.05 across eight measures. Coefficients are median absolute deviation (MAD)-standardized only for cross-measure visualization.

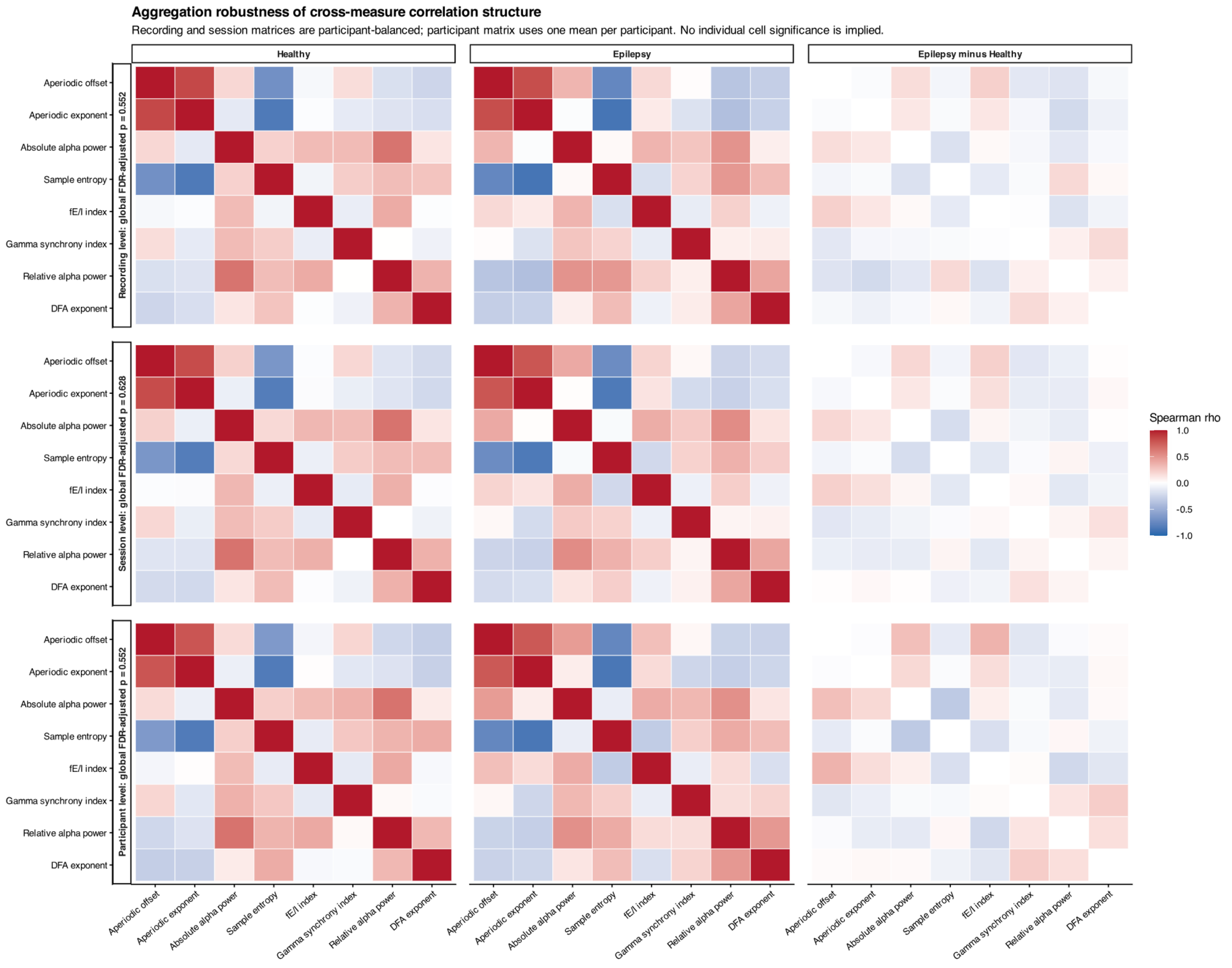

**Figure S2. Aggregation robustness of the cross-measure correlation structure.** Rows show recording-, session-, and participant-level analyses. Recording- and session-level matrices were participant-balanced; the participant-level analysis used one mean per participant. None of the three global group comparisons survived FDR correction across levels. Individual heatmap cells are descriptive and do not represent individual significance tests.

**Table S4. Epilepsy-only ASM-count mixed-effects models**

| Measure | Within person estimate | 95% CI | Raw p | FDR p |
| --- | --- | --- | --- | --- |
| Aperiodic offset | -0.058 | -0.11 to -0.006 | 0.030 | 0.065 |
| Aperiodic exponent | -0.039 | -0.091 to 0.012 | 0.135 | 0.216 |
| Absolute alpha power | 2.26e-13 | -2.57e-13 to 7.09e-13 | 0.358 | 0.477 |
| Sample entropy | 0.04 | 0.003 to 0.076 | 0.033 | 0.065 |
| fE/I index | -0.002 | -0.018 to 0.015 | 0.849 | 0.849 |
| Gamma synchrony index | 0.004 | -0.014 to 0.022 | 0.687 | 0.785 |
| Relative alpha power | 0.008 | 0.004 to 0.012 | <0.001 | 0.001 |
| DFA exponent | 0.008 | 0.002 to 0.014 | 0.015 | 0.059 |

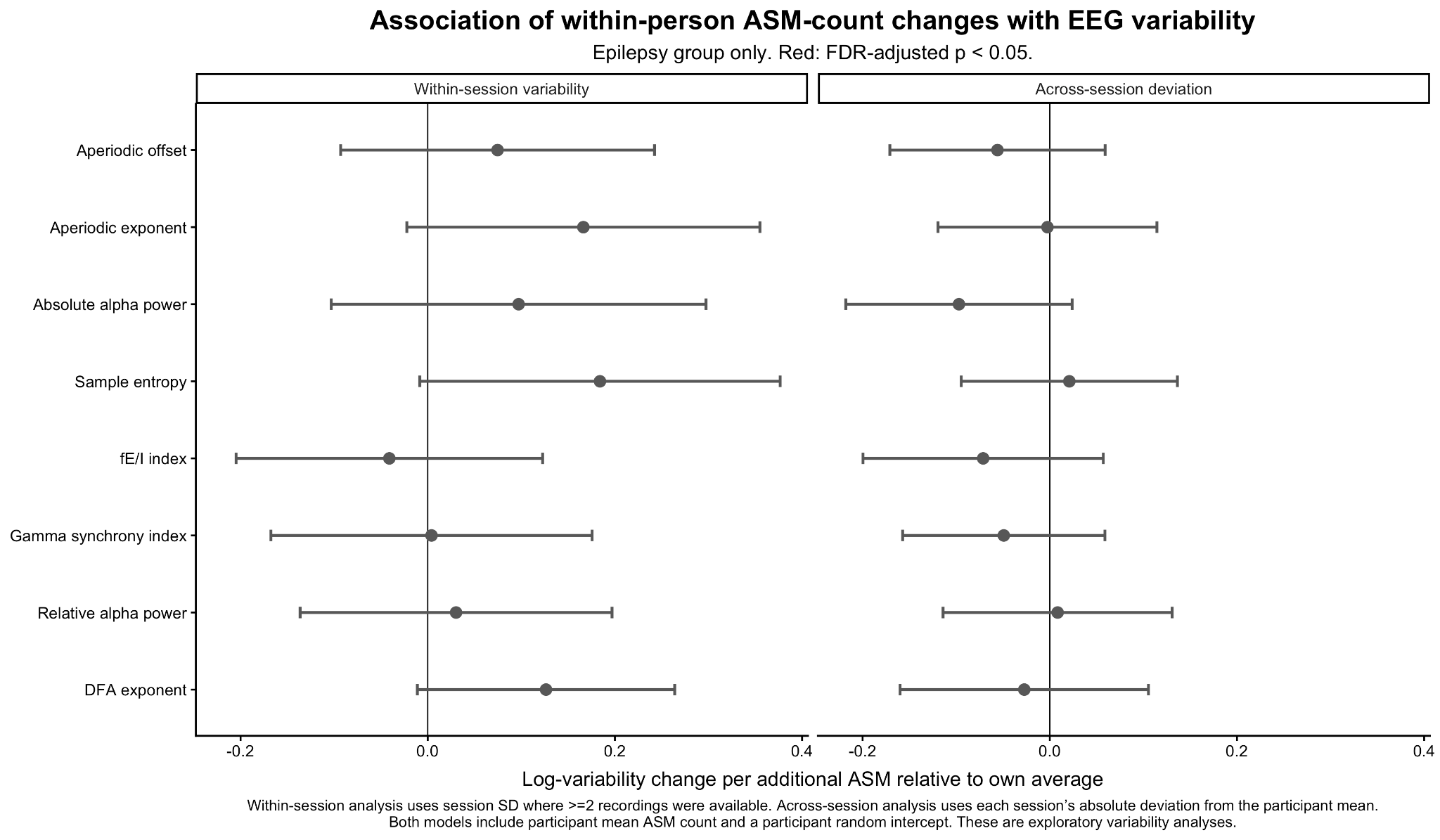

**Figure S3. Exploratory associations of within-person ASM-count change with within-session variability and across-session deviation**. A positive coefficient indicates higher variability with a higher ASM count relative to the participant’s own average. No effect was FDR-significant.

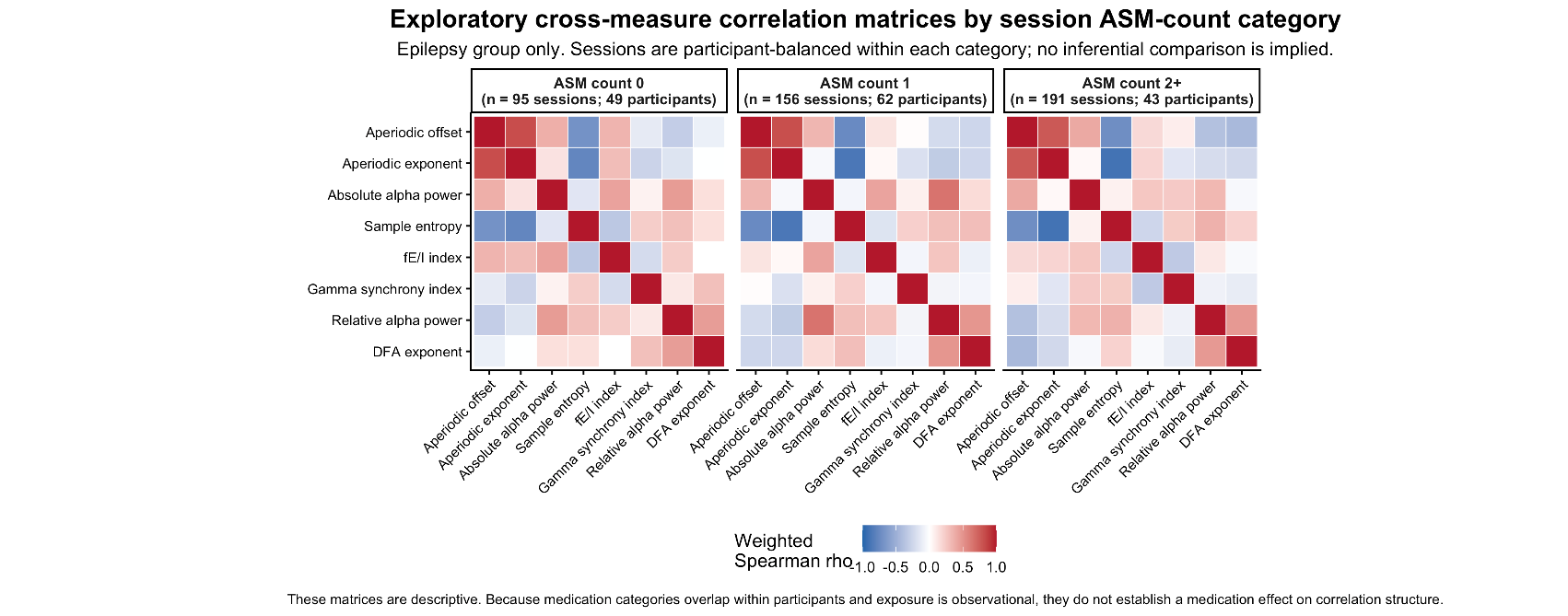

**Figure S4. Descriptive participant-balanced Spearman correlation matrices by session ASM-count category in the epilepsy group.** These displays are not inferential tests of medication effects on correlation structure.
